# Estimates of the rate of infection and asymptomatic COVID-19 disease in a population sample from SE England

**DOI:** 10.1101/2020.07.29.20162701

**Authors:** Philippa M. Wells, Katie J. Doores, Simon Couvreur, Rocio Martinez Nunez, Jeffrey Seow, Carl Graham, Sam Acors, Neophytos Kouphou, Stuart J.D. Neil, Richard S. Tedder, Pedro M. Matos, Kate Poulton, Maria Jose Lista, Ruth E. Dickenson, Helin Sertkaya, Thomas J.A. Maguire, Edward J. Scourfield, Ruth C. E. Bowyer, Deborah Hart, Aoife O’Bryne, Kathyrn J.A. Steel, Oliver Hemmings, Carolina Rosadas, Myra O. McClure, Joan Capedevilla-pujol, Jonathan Wolf, Sebastien Ourselin, Matthew A. Brown, Michael H. Malim, Tim Spector, Claire J. Steves

## Abstract

**Background:** Understanding of the true asymptomatic rate of infection of SARS-CoV-2 is currently limited, as is understanding of the population-based seroprevalence after the first wave of COVID-19 within the UK. The majority of data thus far come from hospitalised patients, with little focus on general population cases, or their symptoms.

**Methods:** We undertook enzyme linked immunosorbent assay characterisation of IgM and IgG responses against SARS-CoV-2 spike glycoprotein and nucleocapsid protein of 431 unselected general-population participants of the TwinsUK cohort from South-East England, aged 19-86 (median age 48; 85% female). 382 participants completed prospective logging of 14 COVID-19 related symptoms via the COVID Symptom Study App, allowing consideration of serology alongside individual symptoms, and a predictive algorithm for estimated COVID-19 previously modelled on PCR positive individuals from a dataset of over 2 million.

**Findings:** We demonstrated a seroprevalence of 12% (51participants of 431). Of 48 seropositive individuals with full symptom data, nine (19%) were fully asymptomatic, and 16 (27%) were asymptomatic for core COVID-19 symptoms: fever, cough or anosmia. Specificity of anosmia for seropositivity was 95%, compared to 88% for fever cough and anosmia combined. 34 individuals in the cohort were predicted to be Covid-19 positive using the App algorithm, and of those, 18 (52%) were seropositive.

**Interpretation:** Seroprevalence amongst adults from London and South-East England was 12%, and 19% of seropositive individuals with prospective symptom logging were fully asymptomatic throughout the study. Anosmia demonstrated the highest symptom specificity for SARS-CoV-2 antibody response.

**Funding:** NIHR BRC, CDRF, ZOE global LTD, RST-UKRI/MRC

## Introduction

The COVID-19 pandemic has constituted an international emergency, accounting world-wide for greater than 600,000 deaths thus far. In order to understand the transmission of the virus through the population, and to estimate protection afforded to those post-infection, we must first understand the seroprevalence to SARS-CoV-2 with regards to demographics and clinical presentation.

Data on the rates of infection in the United Kingdom (UK) come mainly from the Office for National Statistics (Government ONS) surveys. As of 29^th^ June 2020, the ONS estimate 6.3% (95% CI 5 to 7.8) of the UK population to be seropositive for IgG or IgM S glycoprotein detected using enzyme linked immunosorbant assay (ELISA) testing, based on blood tests of 885 individuals since 26 April 2020.^1^,^2^ In the 7^th^ July report from the ongoing ONS household survey involving PCR swab testing of around 20,000 people, the authors reported an asymptomatic infection rate of 66%.^3^

However, these surveys did not study the full range of symptoms associated with COVID-19, and only assessed symptomatology at the time of swabbing, and therefore may have over- estimated this rate. We undertook a population-based study of the humoral immune response to SARS-CoV-2, with regards to longitudinal clinical symptoms collected through a mobile phone app in a population-based sample of 431 TwinsUK volunteers.^2^

## Methods

### Study design and participants

Participants were members of the TwinsUK cohort, the largest UK registry of adult twins.^4^Participants were visited in their home to obtain saliva and serum samples to test for active infection and antibody response. The majority of participants had completed regular logging of symptoms, via the C-19 Covid Symptom Study app^5^since, enabling measurement of antibody response to COVID-19 with regards to clinical symptoms. Participants were members of the TwinsUK cohort, the largest UK registry of adult twins.^4^Participants were visited in their home to obtain saliva and serum samples to test for active infection and antibody response. For three months prior to the visit, the majority of participants had completed regular logging of symptoms, via the C-19 Covid Symptom Study app^5^, enabling measurement of antibody response to COVID-19 with regards to clinical symptoms.

The inclusion criteria for the study were residence within an 80-mile radius of the cohort headquarters St Thomas’ Hospital in Central London, active use of the COVID symptom study app, and availability for visit between 27^th^ April and 2^nd^ June 2020. The exclusion criterion (for safety reasons) was report of recent symptoms indicating potential COVID-19 at the time of the study or within 14 days prior, which participants were required to confirm via telephone consultation. Ethics approval for the study was granted by NHS North West - Liverpool East Research Ethics Committee (REC reference 19/NW/0187), IRAS ID 258513, and all participants gave written informed consent. 895 twins met the geographical and logging based inclusion criteria. Of these, 231 could not be contacted, 154 declined a visit as they were either not interested or did not want a home visit due to being in the high risk category for COVID-19, or because they or a member of their household were currently experiencing symptoms suggestive of COVID-19 infection and were not able or willing to be seen later (n = 22). A further 85 could not be visited within the study period.

Visited participants comprised 431 TwinsUK volunteers, aged between 19 and 86, of whom 367 (85%) were female. They were visited in their home for antibody testing between 27th April and 2nd June 2020, an average of 18 days from the first peak of the pandemic in the UK on the 3rd April 2020. The demographics of the study participants in the context of the wider geographical area in which they reside are summarized in **Table 1**. National data was derived from government published statistics.^6–8^

**Table 1.**
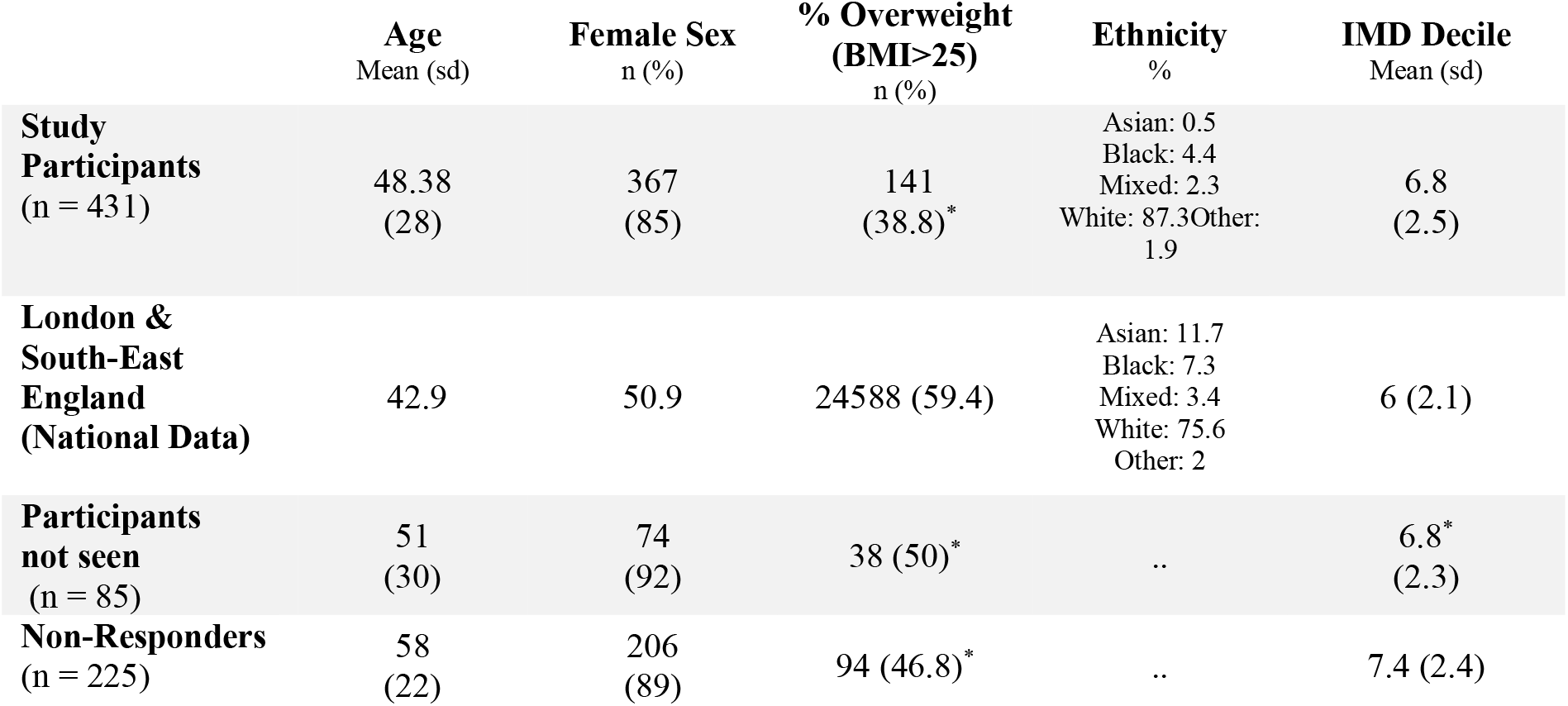
TwinsUK participant demographics.

|  | Age<br>Mean (sd) | Female Sex<br>n (%) | % Overweight<br>(BMI>25)<br>n (%) | Ethnicity<br>% | IMD Decile<br>Mean (sd) |
| --- | --- | --- | --- | --- | --- |
| **Study Participants**<br>(n = 431) | 48.38<br>(28) | 367<br>(85) | 141<br>(38.8)* | Asian: 0.5<br>Black: 4.4<br>Mixed: 2.3<br>White: 87.3<br>Other: 1.9 | 6.8<br>(2.5) |
| **London & South-East England (National Data)** | 42.9 | 50.9 | 24588 (59.4) | Asian: 11.7<br>Black: 7.3<br>Mixed: 3.4<br>White: 75.6<br>Other: 2 | 6 (2.1) |
| **Participants not seen**<br>(n = 85) | 51<br>(30) | 74<br>(92) | 38 (50)* | .. | 6.8*<br>(2.3) |
| **Non-Responders**<br>(n = 225) | 58<br>(22) | 206<br>(89) | 94 (46.8)* | .. | 7.4 (2.4) |

### ELISA testing for antibody response

Serum samples obtained on home visits were tested for IgG and IgM binding to SARS-CoV- 2 S and N proteins using enzyme linked immunosorbent assay (ELISA) using serum diluted at 1:50. The ELISA serology methods used in this study have been previously described.^9^ Briefly, the N/S ELISA demonstrated 100 % specificity (as determined using 300 pre-COVID-19 serum samples), and sensitivity was secondary to time post infection, improving in performance with increasing days from initial infection: after 10 days it was 84.7%. after 14 days it was 87.0% and at greater than 20 days it was 96.4% sensitive. A participant was considered seropositive if an IgG response (optical density (OD) value) to both N and S was detected that was 4-fold above the background of the assay. This cut-off is based on the analysis of 300+ pre-COVID-19 serum samples.^9^

The ELISA was validated by a separate laboratory at Imperial College London, who employed a hybrid double antigen bridging assay (DABA) using immobilised S1 and HRP-labelled S1 receptor binding domain (RBD) of known high specificity (>99.9%) to compare antibody reactivity in 50 unselected samples. Of these, 10 were reactive for RBD using the hybrid DABA whereas the N/S ELISA method determined 9 of these to be seropositive. All samples found reactive in the N/S ELISA were also shown to be reactive using the hybrid DABA. Thus, comparing to hybrid DABA, our ELISA showed sensitivity 90% (95% CI 60- 99, and specificity 100% (95% CI 93-100), 95% CIs were calculated using the Clopper Pearson method.^7^

### Testing for active the presence of SARS-CoV-2

Buccal swabs obtained during the home visits were used to test for current SARS-CoV-2 RNA (and by inference active replication) using RT-PCR as previously described by Lista *et al*.^10^ Briefly, nasopharyngeal swab samples were heat inactivated at 70 °C for 30 min and 140 µl were used to extract RNA with the Beckman Coulter RNAdvance Blood kit, ending with elution in 50 µL of water. For qPCR, the US CDC designed primer/probe set were used for the N gene (N1 and N2) and RNAseP with 5 µL of RNA sample and Taqman Fast Virus 1- Step Master Mix (Thermofisher).

### Longitudinal recording of participants’ symptoms

Longitudinal experience of potential COVID-19 symptoms was prospectively logged by participants via the C-19 Covid Symptom Study app, the primary app for self-report of symptoms during the pandemic in the UK.^5^ The C-19 Symptom Study App has been developed by the health science company ZOE in collaboration with King’s College London. Thus far, 4,007,688 participants have logged symptoms, providing invaluable epidemiologic information with regards to the pandemic. The symptoms which app participants were asked to record are listed in **Table 2**.

**Table 2.** Clinical symptoms included in C-19 Covid Symptom Study App. For each App entry, participants complete a tick-box form to report whether they are presently experiencing any of these symptoms.

| General Symptoms | Core COVID-19 Symptoms |
| --- | --- |
| • Fatigue<br>• Muscle pain<br>• Chest pain<br>• Nausea<br>• Headache<br>• Shortness of breath<br>• Abdominal pain<br>• Diarrhoea<br>• Hoarse voice<br>• Skipped meals<br>• Skin welts or swelling of the face or lips<br>• Sores or blisters on feet<br>• Eye soreness or discomfort<br>• Any other symptom | • Cough<br>• Fever<br>• Anosmia |

### Algorithm for predicting prior COVID-19 from symptoms reported

A predictive algorithm for identification of COVID-19 using longitudinal symptoms reported via the C-19 App was used to identify participants who were predicted to have had prior symptomatic infection, using the method recently described by Menni *et al*.^11^ Briefly, the algorithm was developed from symptoms in people testing positive for SARS_CoV_2 using RT-PCR. The formula includes two core symptoms (anosmia and cough), two non-core symptoms (fatigue and skipped meals), in addition to participant age and sex. In addition, participants were asked whether they had experienced any symptoms suggestive of COVID- 19 prior to launch of the App on 24^th^ March 2020.

### Consideration of serology in relation to longitudinal symptoms logs and statistical analysis

382 participants had regularly logged their symptoms prospectively from App launch on 24^th^ March, and also reported on symptoms retrospectively prior to this date. Symptom profiling included core COVID-19 symptoms and general symptoms, in addition to algorithm prediction of prior COVID-19. Participants were delineated according to antibody status in relation to reported symptoms and predicted COVID-19 prior infection. Using the status regarding prior SARS-CoV-2 infection as predicted by the algorithm, we calculated seroprevalence amongst those who were likely to have had COVID-19. In these participants, we investigated association of demographic factors: age, sex, and BMI, with seroprevalence (Student’s T test for difference between groups). Confidence intervals for proportions were calculated using the Clopper Pearson method.^7^ All analyses were performed using the R software environment for statistical computing.

## Results

In our sample of 431 adults aged 19-89 years living in London and South-East England, seroprevalence using our ELISA assay was demonstrated to be 12% (51 participants, 95% CI = 9.1-15.2).

During the study period, beginning 24^th^ March 2020, a total of 382 participants completed longitudinal logging of symptoms via the C-19 COVID Symptom Study App, providing detailed insight into clinical presentation of this community sample. The median number of App entries to record symptoms, per individual, was 51 days (IQR 30 to 64). 48 (12%; 95% CI 9-16) were seropositive, of whom, 9/48 (19%; 95% CI 9-33) were completely asymptomatic, including no prior symptoms before the App launch. 27% reported only non-core symptoms, (i.e. neither fever, persistent cough, nor anosmia; **Table 3**), and would have been reported as asymptomatic in other surveys.

**Table 3.** Symptom pattern in relation to antibody seropositivity. Any symptom indicates participant’s response to the App including whether they had symptoms they thought were COVID before the App was launched. Anosmia was reported prospectively only.

| Symptoms |  | Serology |  | Sensitivity | Specificity | PPV | NPV |
| --- | --- | --- | --- | --- | --- | --- | --- |
|  |  | + | - | % (95%CI) | % (95%CI) | % (95%CI) | % (95%CI) |
| Any App symptoms | + | 39 | 144 | 81(67-91) | 57 (52-62) | 21 (18-24) | 96 (93-98) |
|  | - | 9 | 190 |  |  |  |  |
| Any core symptoms | + | 35 | 50 | 73 (60-85) | 85 (81-89) | 40 (34-47) | 96 (94-97) |
|  | - | 13 | 284 |  |  |  |  |
| Anosmia alone | + | 23 | 18 | 48 (35-63) | 95 (92-97) | 56 (44-68) | 93 (91-95) |
|  | - | 25 | 319 |  |  |  |  |
| Predicted COVID using algorithm | + | 18 | 16 | 37 (26-53) | 95 (93-97) | 53 (39-66) | 92 (90-93) |
|  | - | 30 | 321 |  |  |  |  |

The symptom which most strongly predicted seropositivity was anosmia. Of participants reporting anosmia, 47.9% had detectable IgG to SARS-CoV-2. The specificity of anosmia for seropositivity was 95%, whereas for fever, cough, and anosmia combined was less at 85%. Both were not highly sensitive with (anosmia 48%; all core symptoms together 73%).

There were 34 individuals in the cohort who were predicted to have had a SARS-CoV-2 infection using the App based algorithm. The algorithm correlated better with serology than did core symptoms alone (**Figure 1B**; **Table 3**). Of the 34 individuals with predicted COVID-19, 52% (18) were seropositive (**Table 3**).

**Figure 1.**
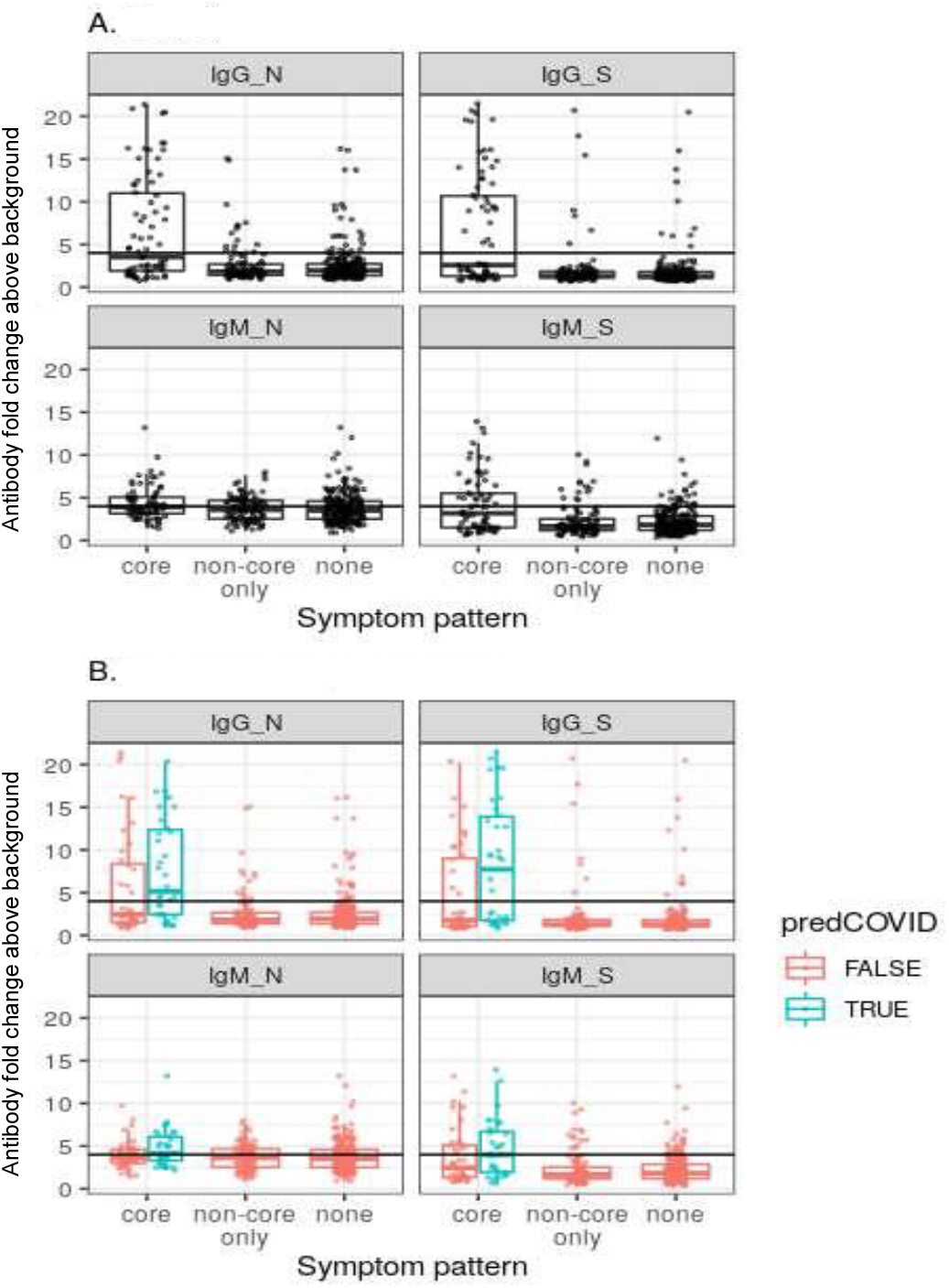
Antibody levels in relation to symptom pattern. Depicted in A) all participants with antibody profiling and in B) participants are partitioned by predicted COVID status (predCOVID), estimated using the algorithm for prior infection of COVID-19, based on reported symptoms. Antibody levels are measured as fold change in absorbance (OD) above test background.

### Factors associated with seroprevalence amongst those with predicted COVID-19

On comparison of the seropositive and seronegative participants with predicted COVID-19 (n = 34, out of 381 participants who prospectively logged their symptoms), seropositive participants were older (median age seropositive 48, median age seronegative 36; p = 0.046). No difference in sex (% female of seropositive participants 72, and 87 for seronegative) or BMI (median 23.8 seropositive; 22.8 seronegative) was evident between the groups.

## Discussion

Understanding seroprevalence is important in order to estimate epidemiological spread of COVID-19 through the population, to inform on the potential efficacy of vaccination, and for the concept of herd immunity which may eventually be achieved via a combination of vaccination and convalescence.^12^ We estimate the seroprevalence rate within our sample to be 12%, higher than the ONS estimate for the UK of 6% (studies summarized in **Supplementary Table 1**), but could be compatible given the potentially higher prevalence in London and the South-East and the fact that, unlike the ONS surveys, we only included adults.

Our results indicate that a substantial portion (19%) of those who have detectable antibodies to SARS-CoV-2 were entirely asymptomatic, confirming that even those who have clinically very mild disease with no perceivable symptoms may produce antibodies. This frequency is substantially lower than most other estimates of asymptomatic COVID-19, which we believe to be due to our more complete assessment of symptoms over time. Indeed, this may still be an overestimate, as, at the start of the survey during the peak epidemic, we did not ask about rashes, which our data suggest are present in 8% of positive cases.^13^ Our estimate relates to asymptomatic development of specific antibodies, rather than asymptomatic carriage of SARS-CoV-2, as it has been reported that not all who have COVID-19 develop detectable antibodies.^14,15^ In a population study by Solbach *et al*. of 110 German participants who had had PCR confirmed prior COVID-19 and who self-reported symptoms, there were ten asymptomatic individuals (11%). Of these ten asymptomatic cases, four were seropositive, and the remainder had no detectable antibodies in two consecutive analyses.^15^ It is clear that asymptomatic disease holds important implications with regards to transmission, as asymptomatic individuals are unaware of their infected status, and may support a significantly longer period of viral shedding, thereby exacerbating the potency of transmission potential.^16,17^

Without parallel PCR testing alongside symptom tracking, we cannot be certain whether seronegative individuals reporting COVID-19 symptoms either did not develop detectable SARS-CoV-2 antibodies, their antibody levels had declined to undetectable^18^ or whether their symptoms related to another infection or condition. Within the 34 people who were predicted to have had COVID-19 using the App based algorithm, there was approximately a 50/50 likelihood of participants having a detectable antibody response (52% were seropositive, whilst 48% had no detectable antibodies). Our understanding of the human immune response to SARS-CoV-2 continues to improve, and emerging evidence indicates that T cell mediated immunity plays an important role in the immune control of COVID-19.^19,20^ For example, a small community-based study of patients and their household families showed than an unexpected six out of eight family members who had been infected demonstrated a COVID- 19-antigen specific T cell response, but had no detectable antibodies.^21^ Speculatively, therefore, T cell mediated responses may afford efficacious immunity in the absence of a detectable antibody response. In the participants with symptoms predictive of COVID-19 using the algorithm, age differed by antibody response: seropositive individuals being older than those who were seronegative (p = 0.046). This may be secondary to increased shielding of older participants, however it is consistent with previous reports of a positive correlation of IgG COVID-19 antibodies with age.^14,17^ Whether T cell mediated reponses are present or more prominent in younger individuals is part of an ongoing follow-on of this study.

Using longitudinal symptoms logged using the App, we demonstrated that of all the individual clinical symptoms, anosmia was the strongest indicator of seropositivity; the specificity of anosmia was 95% and sensitivity was 48 %, whilst the specificity of all core symptoms combined was 85%. Using data from the same Covid Symptom Study App, our group was able to demonstrate previously that anosmia is a core symptom of COVID-19, in addition to the prior recognised symptoms of fever and persistent cough.^8,13^. These results further highlight anosmia as an important core COVID-19 symptom which correlates strongly with both swab positivity and antibody response.

This study, like many similar surveys has a number of limitations. An overarching consideration with regards to seroprevalence to SARS-CoV-2 is the longevity of seropositivity. In a longitudinal study of RT-PCR confirmed prior COVID-19 cases by Doores *et al*., individuals mounted a range of antibody responses, and a decline in levels and virus neutralisation was typically observable within three months of the onset of symptoms.^18^Thus, it is plausible that antibody responses would have fallen below the level of detection by the time of assessment in some of our study participants. Estimation of prior COVID-19 infection was via a symptom based algorithm as PCR confirmation had not been undertaken, nevertheless, we have previously published that this algorithm had a high PPV of close to 80 percent and was trained on symptoms of 7178 swab positive individuals.^11^

No volunteer cohort is fully representative of the general population, but we sampled a range of age and ethnicities and included people from a wide area of deprived and affluent neighbourhoods and with a range of BMI. Despite this, the cohort is detectably more affluent and white,than the general population which would serve to reduce our estimate of prevalence, give the extra burden of disease shouldered by less affluent groups and people of black, asian and minority ethnic background. Additionally we use the index of multiple deprivation which is an area-level indicator rather than individual level socioeconomic- position. The excess of females in our data set reflects the TwinsUK cohort and, so far, differences in seropositivity between genders have been minor in other datasets. It is possible that women report symptoms more readily than men which means that the true assymtomatic rate may be higher. The data are from London and the South East and seroprevalence data from this study will therefore not be generalisable to the whole of the country or to children.

Further work is planned to extend the study using larger numbers from the 4 million app users who reported symptoms to address many of the underlying factors influencing swab positivity and antibody responses. The present study is underpowered in this regard with the asymptomatic seropositive group comprising 9 individuals out of a total of 48 seropositive participants.

In conclusion, we estimated that 12% of the SE England population were seropositive between 27^th^ April and 2^nd^ June, an average of 18 days from the peak of the epidemic in the UK. We estimate the asymptomatic rate to be 19%. Anosmia was the symptom with the highest specificity for seropositivity. These data should be useful for both epidemiology and public health planning and reinforces the need to collect good symptom data as neither PCR testing nor antibody tests adequately capture all disease.

## Data Availability

Data is available upon reasonable request to TwinsUK.

## Acknowledgements

This study was supported by a COVID urgent response grant from the Chronic Disease Research Foundation (CDRF). Zoe Global Limited developed the app with the guidance of clinicians. Investigators received support from the Wellcome Trust, the MRC/BHF, Alzheimer’s Society, EU, NIHR, CDRF, RST-UKRI/MRC and the NIHR-funded BioResource, Clinical Research Facility and BRC based at GSTT NHS Foundation Trust in partnership with KCL. We thank the volunteers of TwinsUK without whom this work would not be possible, and all participants who entered data into the C-19 Covid Symptom Study App. We thank the staff of Zoe Global, the Department of Twin Research at King’s College London and the Clinical and Translational Epidemiology Unit at Massachusetts General Hospital for tireless work in contributing to the running of the study and data collection. We thank E. Segal and his laboratory for helpful input. Thank you to Philip Brouwer, Marit Van Gils and Rogier Sanders (University of Amsterdam) for the S protein construct, and Leo James, Jakub Luptak and Leo Kiss (LMB) for the provision of purified N protein. This work was supported by Zoe Global. The Department of Twin Research receives grants from the Wellcome Trust (212904/Z/18/Z) and Medical Research Council/British Heart Foundation Ancestry and Biological Informative Markers for Stratification of Hypertension (AIMHY; MR/M016560/1), and support from the European Union, the Chronic Disease Research Foundation, Zoe Global, the NIHR Clinical Research Facility and the Biomedical Research Centre (based at Guy’s and St Thomas’ NHS Foundation Trust in partnership with King’s College London). King’s Together Rapid COVID-19 Call awards to MHM, KJD, SJDN and RMN. MRC Discovery Award MC/PC/15068 to SJDN, KJD and MHM. RST-UKRI/MRC award to RT (MC_PC_19078). CG and RED were supported by the MRC-KCL Doctoral Training Partnership in Biomedical Sciences (MR/N013700/1). SA was supported by an MRC-KCL Doctoral Training Partnership in Biomedical Sciences Industrial Collaborative Award in Science & Engineering (iCASE) in partnership with Orchard Therapeutics (MR/R015643/1). NK was supported by the Medical Research Council (MR/S023747/1 to MHM). Wellcome Trust (106223/Z/14/Z to MHM). SJDN was supported by a Wellcome Trust Senior Fellowship (WT098049AIA). Fondation Dormeur, Vaduz for funding equipment (KJD). We thank the TwinsUK Operations Team who conducted home visits for the study; Main members: Deborah Hart, Darioush Yarand, Christel Barnetson, Paz Garcia, Alyce Sheedy, Ayrun Nessa, Genevieve Lachance, Gabriela Surdulescu, Andrei Baleanu, Sam Wadge, Gulsah Akdag, and Paul Snook; Ad hoc: Andrew Anastasiou, Rachel Seymour, and Rajan Wignarajah. We thank Bea Zazzara for assisting with the figures.

## Author Contributions

CJS, TS & MHM designed and implemented the study. KJD led methods for serology. JS, CG, SA, AB, KJAS, OH and NH undertook the ELISAs, CR and MoM undertook the DABAs, and PMM, KP, MJL, RED, HS, TJAM and EJS performed the PCR work. SC analysed the ELISA and App data. CJS supervised the analysis. PMW contributed to data analysis and drafted the manuscript. RB curated the demographic data. All authors contributed to reviewing of the manuscript.

**We declare no conflicts of interest**.

## Supplementary Material

**Supplementary Table 1.** Summary of prior studies of the general population antibody response to SARS-COV-2.

| Methods Summary | Key Findings | Reference |
| --- | --- | --- |
| Protein microarray using serum samples. Testing of 7773 individuals in the **USA**.<br>Antigens: S1 glycoprotein, Receptor binding domain (RBD), S2 glycoprotein and SARS-CoV2 using serum samples. | The main purpose of the study was to demonstrate efficacy of their tests. Population antibody responses are not given in detail and specific antigens are not reported.<br>13.1% population sample seropositive for IgM<br>12.3% population sample seropositive for IgG<br>5% population sample seropositive for IgA<br>20.4% population sample seropositive for any antibody. | Krishnamurphy *et al.*<sup>19</sup><br>*Preprint* |
| From 149 COVID-19 convalescent individuals recruited from Rockefeller University Hospital (New York, **USA**), plasmas were collected an average of 39 days after the onset of symptoms, and performance evaluated in SARS_COV_2 pseudovirus neutralizing assay. | Plasma samples had variable half-maximal pseudovirus neutralizing titres: less than 1:50 in 33% and below 1:1,000 in 79%, while only 1% showed titres above 1:5,000. Antibody sequencing revealed expanded clones of RBD-specific memory B cells expressing closely related antibodies in different individuals. Despite low plasma titres, antibodies to three distinct epitopes on RBD neutralized at half-maximal inhibitory concentrations (IC50 values) as low as single digit nanograms per millilitre.<br>Most convalescent plasmas obtained from individuals who recover from COVID-19 do not contain high levels of neutralizing activity. | Robbiani *et al.*<sup>2</sup><br>*(Accelerated preview)* |
| IgA and IgG S antibody profiles from 110 **German** patients with self-reported mild to moderate, or no COVID-19 related symptoms after laboratory-confirmed infection with SARS-CoV-2. | Found that approximately 70 % of patients developed antibodies 3 weeks or later after the infection. In about 30 % of patients with no or mild to moderate symptoms, no significant antibodies could be detected in two consecutive analyses. Of the 10 asymptomatic individuals, four were repeatedly positive and six had no specific antibodies. | Solbach *et al.*<sup>3</sup><br>*Preprint* |
| Investigated using Luminex assay in 578 **Spanish** health care workers, seroprevalence to SARS-COV-2 including IgM, IgG and IgA antibodies to the receptor binding domain (RBD) | Of the 578 total participants, 39 (6.7%, 95% CI: 4.8-9.1) had been previously diagnosed with COVID-19 by rRT-PCR, 14 (2.4%, 95% CI: 1.4-4.3) had a positive RT-PCR at recruitment, and 54 (9.3%, 95% CI: 7.2-12.0) were seropositive for IgM and/or IgG and/or IgA against SARSCoV-2.<br>Of the 54 seropositive health care workers, 21 (38.9%) had not been previously diagnosed with COVID-19, though 10 of these (47.6%) reported past COVID-19-compatible symptoms.<br>The cumulative prevalence of SARS-CoV-2 infection was 11.2% (65/578, 95% CI: 8.9-14.1). Among those with evidence of past or current infection, 46.2% (12/26) had history of COVID-19-compatible symptoms. IgM levels positively correlated with age ($\rho=0.36$, $p$-value=0.031) and were higher in participants with more than 10 days since onset of symptoms ($p$-value=0.022), and IgA levels were higher in symptomatic than asymptomatic subjects ($p$-value=0.041). | Garcia-Basteiro *et al.*<sup>5</sup><br>*Preprint* |

**Supplementary Figure 1.**
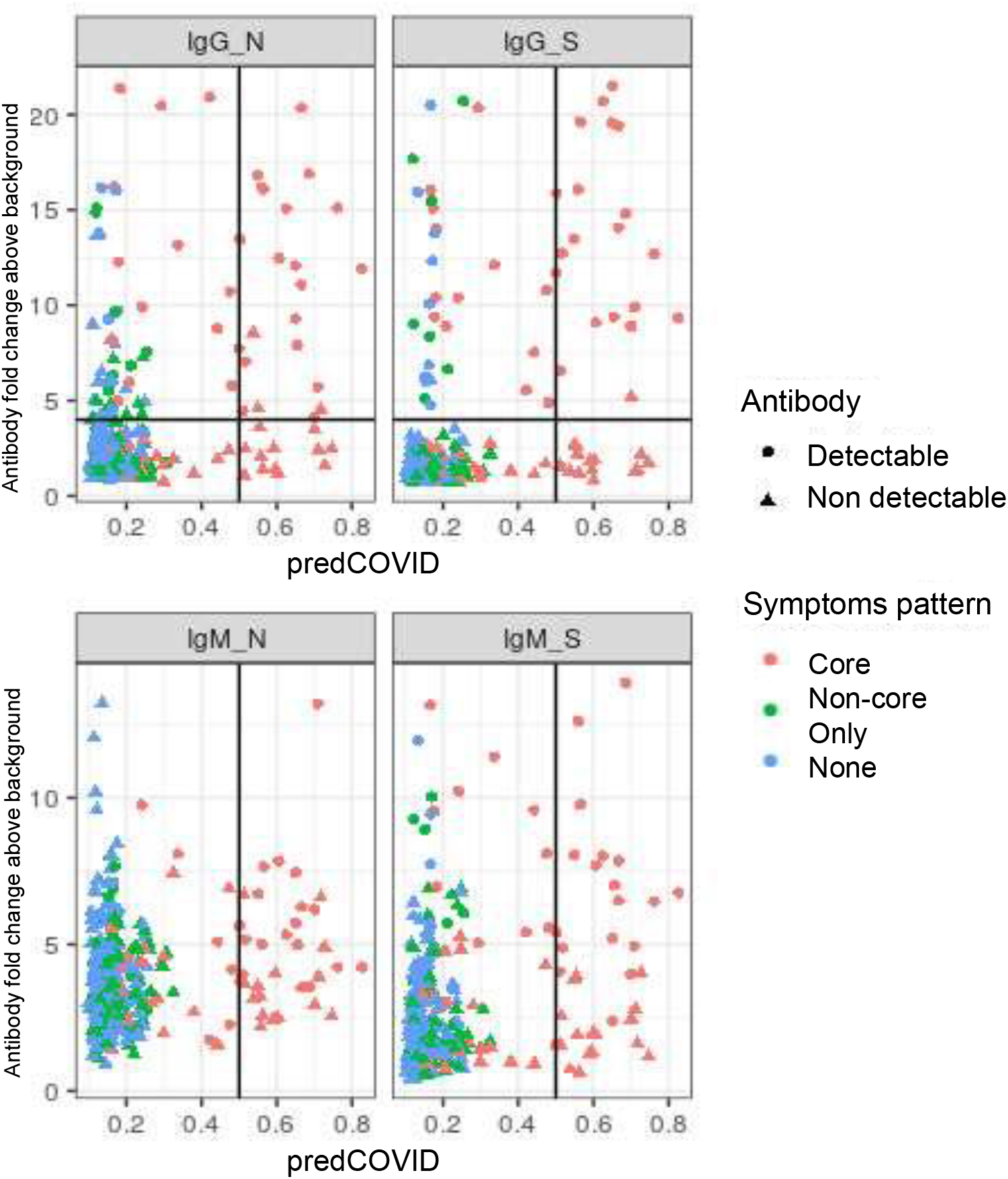
Antibody response in relation to symptom pattern. Core symptoms are defined as fever, cough and/or anosmia. Colour depicts symptom pattern of core symptoms (red), non-core symptoms (green), and asymptomatic (blue). Shape depicts antibody status of detectable (positive; circle) or undetectable (negative; triangle). Antibody level denotes fold change above background, and a threshold of <4 indicates seropositivity. Predicted COVID-19 (predCOVID) is the algorithm score predicting which participants are likely to have had COVID-19 using symptoms reported via the C-19 App. The threshold for predicted COVID-19 of 0.5 is marked.

Visualisation of longitudinal symptom reporting using heat maps demonstrated that anosmia clearly delineated seropositive from seronegative individuals (**Supplementary Figure 2**).

**Supplementary Figure 2.**
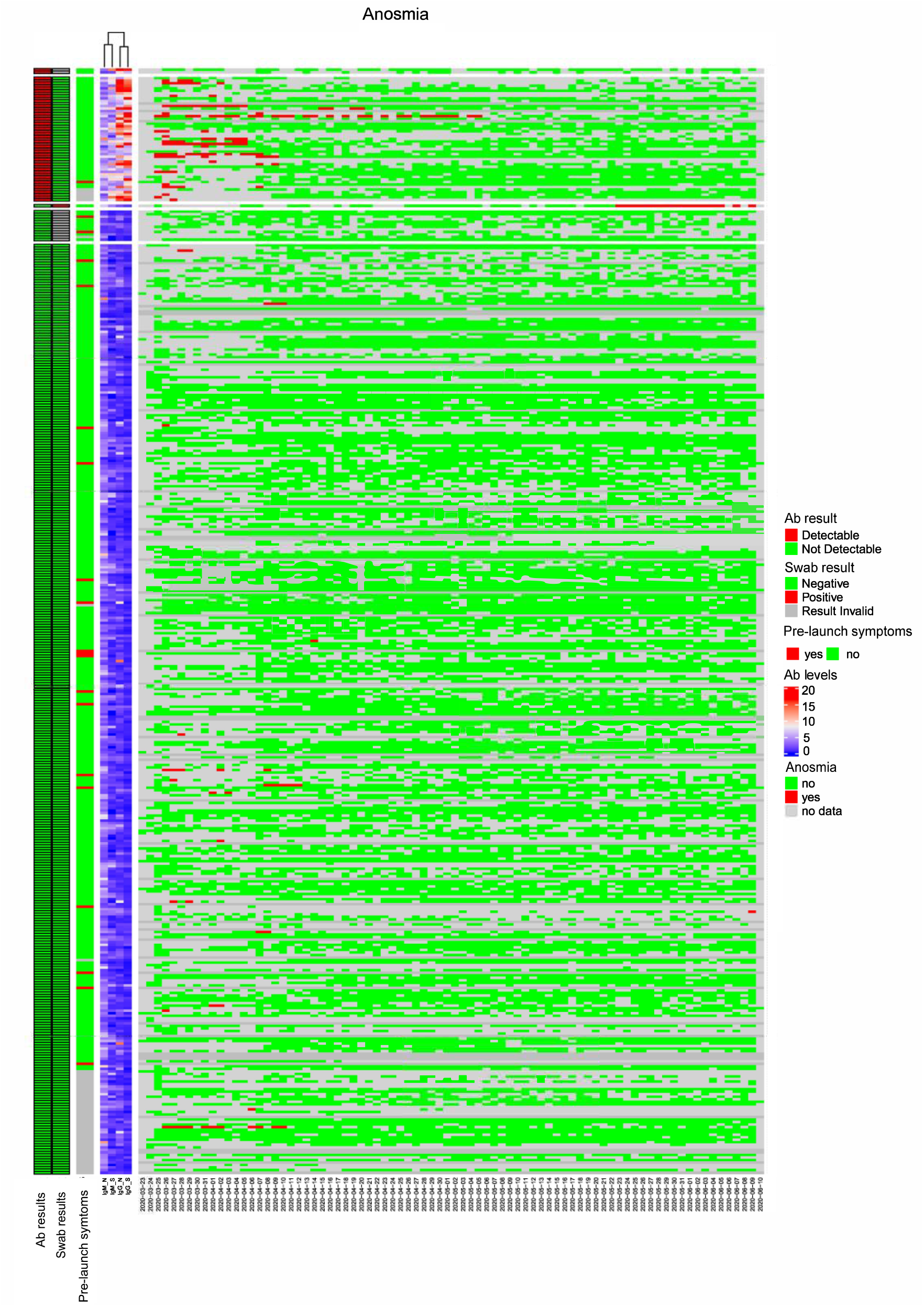
Heatmap of longitudinal anosmia symptom reporting partitioned by seropositivity for antibodies to SARS-COV-2. Samples are clustered according to levels of antibodies for IgG S protein, recorded as fold change above test background. Result of PCR testing of buccal swab samples for current infection is indicated (red indicates positive). Longitudinal reporting of anosmia is given per day, with red indicating a positive answer. The column labelled “Prelaunch symptoms” indicates where a participant reported they had already had COVID symptoms before 24th March.

